# Whole-genome analysis of monozygotic twins discordant for type 1 narcolepsy

**DOI:** 10.1101/2021.08.23.21262137

**Authors:** João H. Campos, Ana C. R. Aguilar, Fernando Antoneli, Giselle Truzzi, Marcelo R. S. Briones, Renata C. Ferreira, Fernando M. S. Coelho

## Abstract

Narcolepsy type 1 (NT1) is a rare and chronic neurological disease characterized by sudden sleep attacks, overwhelming daytime drowsiness, and cataplexy. To contribute to the understanding of NT1 genetic causes, here we describe a whole-genome analysis of a monozygotic twin pair discordant for NT1. Our study revealed that although both twins have the same pathogenic mutations in NT1 associated genes (such as HLA-DQB1*06:02:01, HLA-DRB1*11:01:02/*15:03:01) the unaffected twin has mutations in genes outside the HLA loci that could be suppressing the NT1 phenotype. These results support the notion that NT1 has an immunological basis but that protective mutations in non-HLA might interfere with the clinical manifestation of the disease.

## Introduction

Narcolepsy is a rare neurological disease. Individuals affected by narcolepsy find it difficult to stay awake for long periods, causing serious disruptions in their daily routine. When associated with a sudden loss of muscle tone (cataplexy) narcolepsy is classified as type 1, while the absence of cataplexy indicates type 2 narcolepsy. Genetic, degenerative, and immunological hypotheses try to explain the pathophysiology of narcolepsy. Recently, several findings strongly suggest immunological causes for excessive daytime sleepiness, cataplexy, and other REM disturbances^1^. In narcolepsy type 1 (NT1), the higher prevalence of allele HLA-DQB1*0602 and Hypocretin (Hcrt) deficiency, supports the immunological theory for the cause of NT1^2^. Genes that affect the CD4+ T cells antigen response influence the interaction of DQB1*06:02 and specific T cell receptors (TCRs)^3^. The autoantigen Trib2 is involved in human narcolepsy with anti-Tribble antibodies acting on hypocretin-producing neurons, leading to their disappearance and consequent Hcrt deficiency^4^. It has been proposed that a process of molecular mimicry must be involved given the observed increase of NT1 prevalence after the H1N1 influenza pandemic in 2009–2010^5^.

Excessive daytime sleepiness and cataplexy are keys for the diagnosis of narcolepsy, although many comorbidities are frequently observed in narcolepsy patients. In a study of a narcolepsy community sample, it has been observed a higher prevalence of obstructive sleep apnea, chronic low back pain, obesity, depression, and hyposmia^6^. Narcolepsy is related to other diseases such as Neuromyelitis Optica and thyroid disease with a possible overlap of autoimmune mechanisms^7^. Current findings on narcolepsy are the result of clinical, genetic, and laboratory studies of several research groups worldwide^8,9^. Although discordant monozygotic twins for NT1 are very rare, a previous study characterized the HLA alleles in a twin pair discordant for NT1 and multiple sclerosis^10^. In the present study we report the first whole genome-analysis of monozygotic twins discordant for NT1 in the literature.

## Methods

### Ethics Committee Approval

The ethics protocols for this study were approved by the *Committee on Research Involving Humans* of the Federal University of São Paulo, Brazil. The research team provided the proper written informed consent forms to study participants.

### Participants

This study involved a male pair of late teens, monozygotic twins discordant for narcolepsy type I, that have been admitted at the Daytime Excessive Sleepiness Ambulatory of the Federal University of Sao Paulo – UNIFESP.

The twin with narcolepsy had excessive daytime sleepiness (Epworth Sleepiness Scale (ESS) of 20), cataplexy, hypnagogic hallucinations, polysomnography without abnormalities, multiple sleep latency tests (MSLT) positive for narcolepsy (a mean sleep latency of 3 minutes and sleep-onset REM periods (SOREMPs) of 5), presence of allele HLA-DQB1*0602, and Hypocretin-1 level of zero pg/mL. He was treated with methylphenidate (20 mg/day) and imipramine (25 mg/day), with an improvement of symptomatology.

The other twin had no narcolepsy symptoms (ESS of 4), normal polysomnography, MSLT without abnormalities, presence of allele HLA-DQB1*0602, and Hypocretin-1 level of 396,74 pg/mL^11,12^.

### Whole-Genome Sequencing

After signing the Informed Consent by the guardian and the Informed Consent Terms by both research participants, blood samples were collected. DNA samples were extracted using the QIAamp DNA Blood Mini Kit (Qiagen) according to the manufacturer’s instructions and then sent for DNA sequencing at Centogene AG (Rostock, Mecklenburg-Vorpommern, Germany). Next-generation sequencing (NGS) of whole genomes was performed using Illumina HiSeq, with an approximately 30-fold average read depth of coverage.

### Genome Assembly

The pre-processing of FASTQ files which comprises quality control, adapter trimming, and quality filtering of raw data was done with fastp with recommended parameters^13^. All good reads were then aligned based on the human genome GRCh38 reference (https://www.ncbi.nlm.nih.gov/grc/human) using the Burrows-Wheeler transformation method, through BWA-MEM algorithm (bwa version 0.7.17)^14^ After the initial mapping, resulting BAM files were submitted to the GATK best practices for rigid quality control of assemblies^15^.

### Variant Identification

Variant calling step was done using the GATK’s HaplotypeCaller algorithm, DeepVariant^16^ or Illumina’s Strelka2^17^ tool. When HaplotypeCaller was used, our filtering criteria followed the adjusting parameters as described elsewhere^18^. Strelka2 and DeepVariant were used with recommended default settings. The variant call consensus made by three tools (GATK, Strelka2, and DeepVariant) in each of the twins were obtained using RTG tools (version 3.10-5604f7a) (vcfeval algorithm)^19^. Unique variants were also found with vcfeval. When necessary, visual inspection of the assemblies was employed to increase the confidence in calls and to reduce the risk of false positives using IGV^20^. Finally, the annotation step of genetic variants was conducted using The Ensembl Variant Effect Predictor (VEP), API version 103^21^.

### Haplogroup Identification

Mitochondrial haplogroup profiles were identified using HaploGrep2^22^.

### HLA Typing

To identify HLA alleles, DNA samples were extracted using the QIAamp DNA Blood Mini Kit (Qiagen) according to the manufacturer’s instructions and sent for DNA sequencing at Instituto de Imunogenética IGEN/AFIP (São Paulo, São Paulo State, Brazil). NGS of HLA genes were obtained using Illumina MiSeq, with an approximately 1000-fold average read depth of coverage. TypeStream™ Visual NGS Analysis Software (One Lambda, Canoga Park, California) was used to determine HLA typing.

### Variant Prioritization

To prioritize DVMTs with potential clinical relevance, 3 main filtering criteria were established: (I) being a non-disruptive variant that might change protein effectiveness (impact of consequences predicted as “moderate”), (II) being variant assumed to have a high (disruptive) impact in the protein, probably causing protein truncation, loss of function or triggering nonsense-mediated decay (impact of consequences predicted as “high”), or (III) being considered damaging or deleterious variant by any of the 2 algorithms, SIFT or PolyPhen (included in VEP annotation output). We additionally evaluated DVMTs located in genes associated with NT1 (reported in the literature), or which have clinical data in ClinVar database https://www.ncbi.nlm.nih.gov/clinvar/.

### Overrepresentation Analysis

Genes containing prioritized DVMTs were evaluated into hypergeometric tests using Gene Ontology (GO) data sets for biological processes (BP), cellular components (CC), or molecular functions (MF) with GENE2FUNC tool^23^.

### Smell Identification Test

The University of Pennsylvania Smell Identification Test (UPSIT) is a 40-item olfactory identification test, defining the olfaction acuity as “anosmia”, “severe hyposmia”, “moderate hyposmia”, “mild hyposmia”, and “normal”^24^.

## Results

The analysis strategy used to find divergent variations in monozygotic twins (DVMTs) is outlined in **Figure 1**, and a summary of the annotation of DVMTs found is represented in **Figure 2**. The comparison with the human genome reference (GRCh38), revealed 5,257,409 variants in the unaffected twin, and 5,310,227 variants in the affected. A total of 5,153,368 variants were common to twins. 104,041 variants were unique to the unaffected twin, and 156,863 were unique to the affected one. Unique variants of each twin were classified as DVMTs (total = 260,904) **(Figures 1 and 2)**.

**Figure 1.**
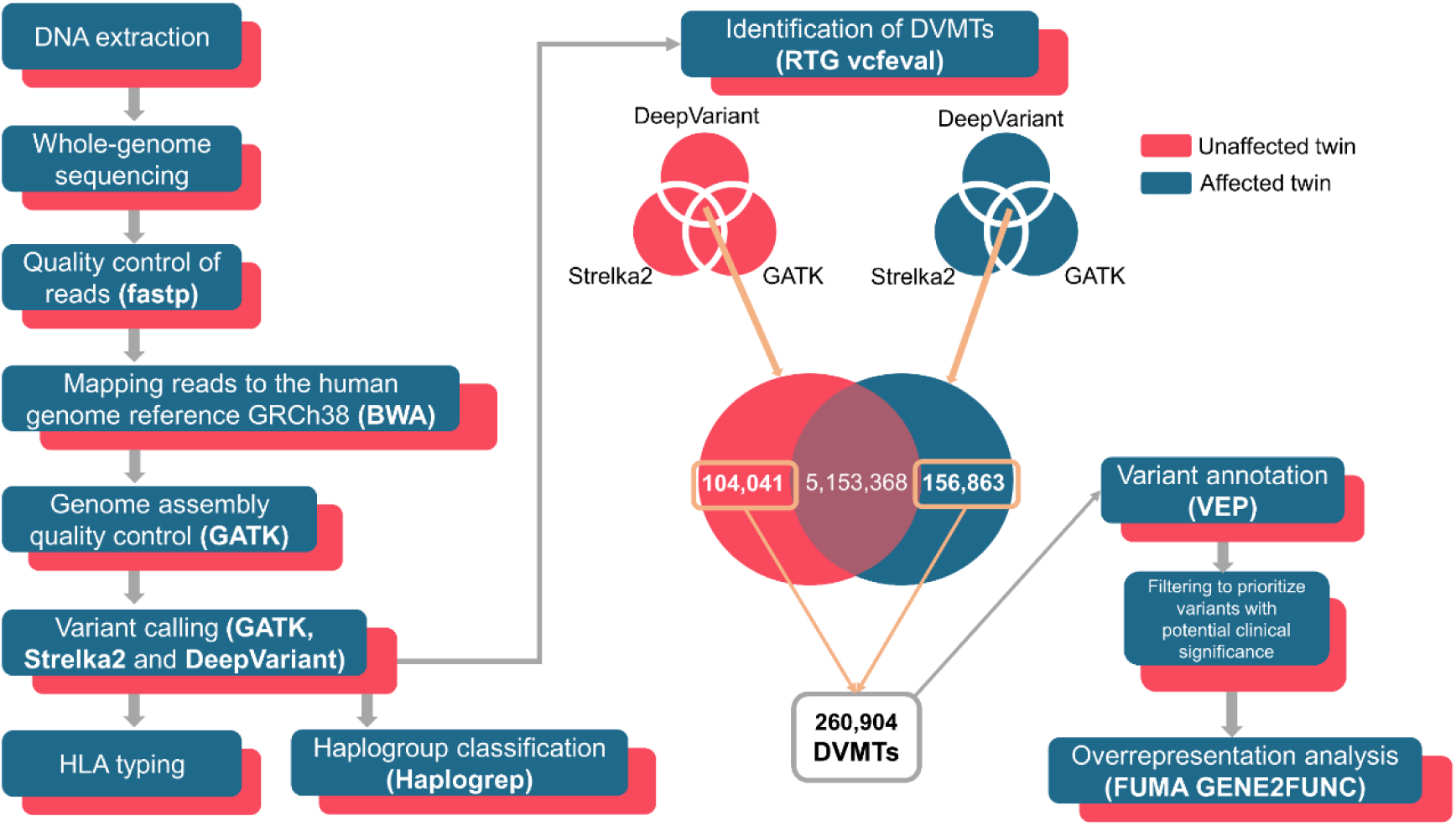
Genomic analysis pipeline of discordant twins for type 1 narcolepsy. Genomes of discordant twins for type 1 narcolepsy were sequenced, and the resulting reads processed for quality control. Good quality reads were then mapped against the human genome reference GRCh38. A new quality control over each assembly was carried out, and the variant calling step was conducted using three independent tools. After that, the consensus between variant call was determined for each twin: in the unaffected one, this resulted in a total of 5,257,409 variants, whereas in the affected twin, 5,310,227 variants. Unique variants were: 104,041 variants in the unaffected twin, and 156,863 variants in the affected twin, and classified as divergent variations in monozygotic twins (DVMTs). Only these 260,904 DVMTs were functionally annotated and filtered to prioritize SNVs with potential clinical relevance. Non-disruptive variants that might change protein effectiveness, or variants assumed to have high (disruptive) impact in the protein, probably causing protein truncation, loss of function or triggering nonsense mediated decay were included in a further enrichment analysis to provide insights about biological processes, cellular components and molecular functions of these DVMTs. Additionally, mitochondrial haplogroups, as well as the typing for the HLA alleles of each twin were determined. The parentheses highlight the tools used for each step.

**Figure 2.**
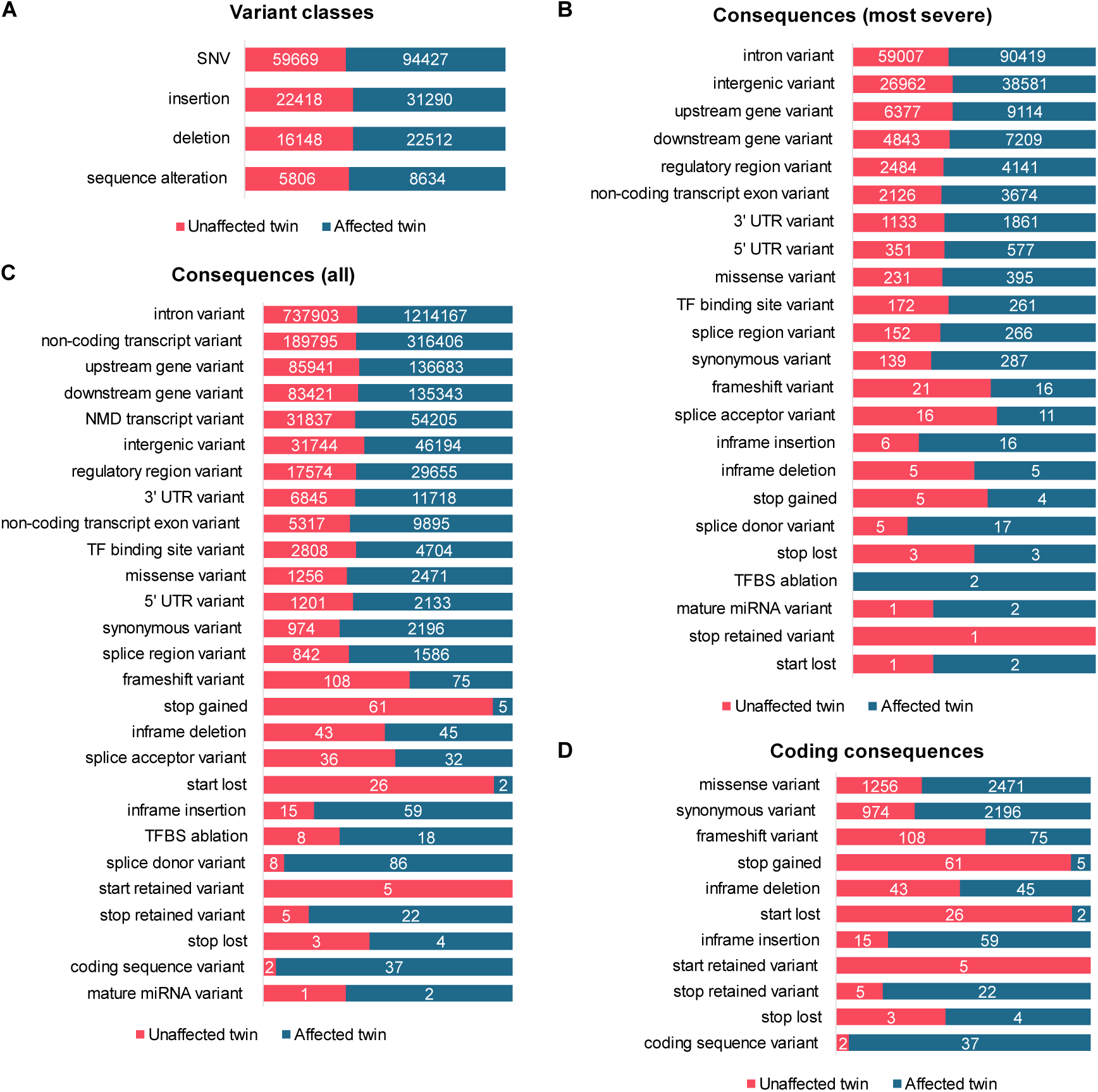
Annotation summary of divergent variations in monozygotic twins (DVMTs). All DVMTs were summarized for variant classes **(A)**, most severe consequences **(B)**, all consequences **(C)**, and for coding consequences **(D)**. Altogether, 104,041 DVMTs were found in the unaffected twin, and 156,863 in the affected twin (total = 206,904). In all summaries, the absolute count for each rating is indicated inside the bars (bars are proportional to the percentages found).

In the unaffected twin, 244 loci seen have DVMTs with the moderate impact of consequence (232 missense variants, 6 inframe insertions, 6 inframe deletions). These variants are distributed over 186 genes, most often in *MUC3A, MUC17*, and *HLA-B* **(Supplementary Table S1)**. As for the variants with the impact of consequence classified as high, these are distributed in 51 loci (24 frameshift variants, 16 splice acceptor variants, 5 splice donor variants, 5 stop gained variants, 3 stop lost variants and 2 start lost variants – the number of consequences is greater because each variant can affect more than one transcript) occurring in 48 genes, mainly in *HGC6*.*3, HLA-A* and *HLA-B* **(Supplementary Table S2)**. 68 variants were classified as deleterious by the SIFT predictor, mainly in the genes *MUC17, MUC12*, and *GOLGA6L2* **(Supplementary Table S3)**. The PolyPhen predictor, on the other hand, identified as damaging, 43 variants, in 37 genes (also mostly in *MUC17, MUC12*, and, additionally, in *FOXO3B*) **(Supplementary Table S4)**. Damaging or deleterious variants identified with the two predictors add up to 29 and are located over 24 genes, occurring more frequently in *MUC12, FOXO3B*, and *MAP2K3* **(Supplementary Table S5)**. 147 loci have variants in 37 genes associated with NT1 (described in the literature), notably in frequency in *HLA-DRB1, CACNA1C, HLA-B, NFATC2*, and *CCR3* **(Supplementary Table S6)**. Among these genes, only *HLA-B* has variants with moderate or high impact of consequences (7 in total, where 5 are missense, and 2 are frameshift variants) **(Supplementary Table S7)**. 103 variants were identified having clinical data in the ClinVar database, of which 13 have uncertain significance or conflicting interpretations of pathogenicity for 23 traits **(Supplementary Table S8)**.

In the affected twin, 421 loci have DVMTs with moderate impact (399 missense, 16 inframe insertions, 8 inframe deletions), distributed in 323 genes, most often in *MUC4, LOC107986175, MUC20, MUC3A*, and *HLA-C* **(Supplementary Table S9)**. As for the high impact variants, these are distributed in 53 loci (17 splice donor variants, 16 frameshift variants, 11 splice acceptor variants, 4 stop gained variants, 3 stop lost variants, and 2 start lost variants) occurring in 50 genes, mainly in *UBXN11, HLA-B*, and *PEX5* **(Supplementary Table S10)**. 99 variants were classified as deleterious by the SIFT, mainly in the genes *MUC12, MUC4*, and *CLCN2* **(Supplementary Table S11)**. PolyPhen identified 66 damaging variants in 55 genes (mostly in *MUC4, HLA-A*, and *HTR3D*) **(Supplementary Table S12)**. Damaging or deleterious variants identified with the two predictors total 46, arranged in 42 genes, and more frequently in *HLA-A, MUC20*, and *MUC4* **(Supplementary Table S13)**. 177 loci have variants in 31 genes associated with NT1, notably in *HLA-B, CACNA1C, HLA-DRB1, SCP2*, and *NFATC2* **(Supplementary Table S14)**. Among these, only *HLA-B* has variants with a consequence of moderate or high impact (6 in total, where 4 are missense and 2 are frameshift variants) **(Supplementary Table S15)**. Finally, 162 variants were identified having data in ClinVar, of which 26 have uncertain significance or conflicting interpretations of pathogenicity for 31 traits **(Supplementary Table S16)**.

As for HLA typing, the twins have the same results for all tested HLA genes **(Table 1**). For genes *HLA-A, HLA-B* and *HLA-C* (MHC class I), the alleles are heterozygous; as for the MHC class II genes, only the *HLA-DQA1* and *HLA-DQB1* alleles are homozygous. The mitochondrial haplogroup of the twins is L3e2b +152, associated with Middle Eastern/African ancestry.

**Table 1.** HLA typing of the monozygotic twin pair discordant for NT1.

| HLA alleles | Unaffected | Affected |
| --- | --- | --- |
| *HLA-A* | *\*30:02:01 / \*34:02:01* | *\*30:02:01 / \*34:02:01* |
| *HLA-B* | *\*14:02:01 / \*53:01:01* | *\*14:02:01 / \*53:01:01* |
| *HLA-C* | *\*04:01:01 / \*08:02:01* | *\*04:01:01 / \*08:02:01* |
| *HLA-DPA1* | *\*01:03:01 / \*03:01:01* | *\*01:03:01 / \*03:01:01* |
| *HLA-DPB1* | *\*02:01:02 / \*04:02P* | *\*02:01:02 / \*04:02P* |
| *HLA-DQA1* | *\*01:02:01 / \*01:02:01* | *\*01:02:01 / \*01:02:01* |
| *HLA-DQB1* | *\*06:02:01 / \*06:02:01* | *\*06:02:01 / \*06:02:01* |
| *HLA-DRB1* | *\*11:01:02 / \*15:03:01* | *\*11:01:02 / \*15:03:01* |
| *HLA-DRB3* | *\*02:02:01* | *\*02:02:01* |
| *HLA-DRB5* | *\*01:01:01* | *\*01:01:01* |

Overrepresentation analysis of genes containing prioritized DVMTs of the unaffected twin showed 16 overrepresented in the Biological Properties (BPs) category, 9 in the Cellular Components (CCs) category, and 1 in the Molecular Function (MF) category **(Figure 3)**. BPs such as “antigen processing and presentation (APP) of endogenous peptide antigen via MHC class I via ER pathway” (GO:0002484), “APP of peptide antigen via MHC class Ib” (GO: 0002428), “APP of exogenous peptide antigen via MHC class I, TAP (transporter associated with antigen processing)-independent” (GO:0002480), “APP of endogenous peptide antigen” (GO:0002483), and “APP via MHC class Ib” (GO:0002475) have a proportion of overlapping genes in gene sets between ∼18-45% **(Figure 3A)**. The highest proportion of overlapping genes in gene sets for CC (∼33%) was for “MHC class I protein complex” (GO:0042612) **(Figure 3B)**, and “olfactory receptor activity” for MF (GO:0004984) **(Figure 3C)**.

**Figure 3.**
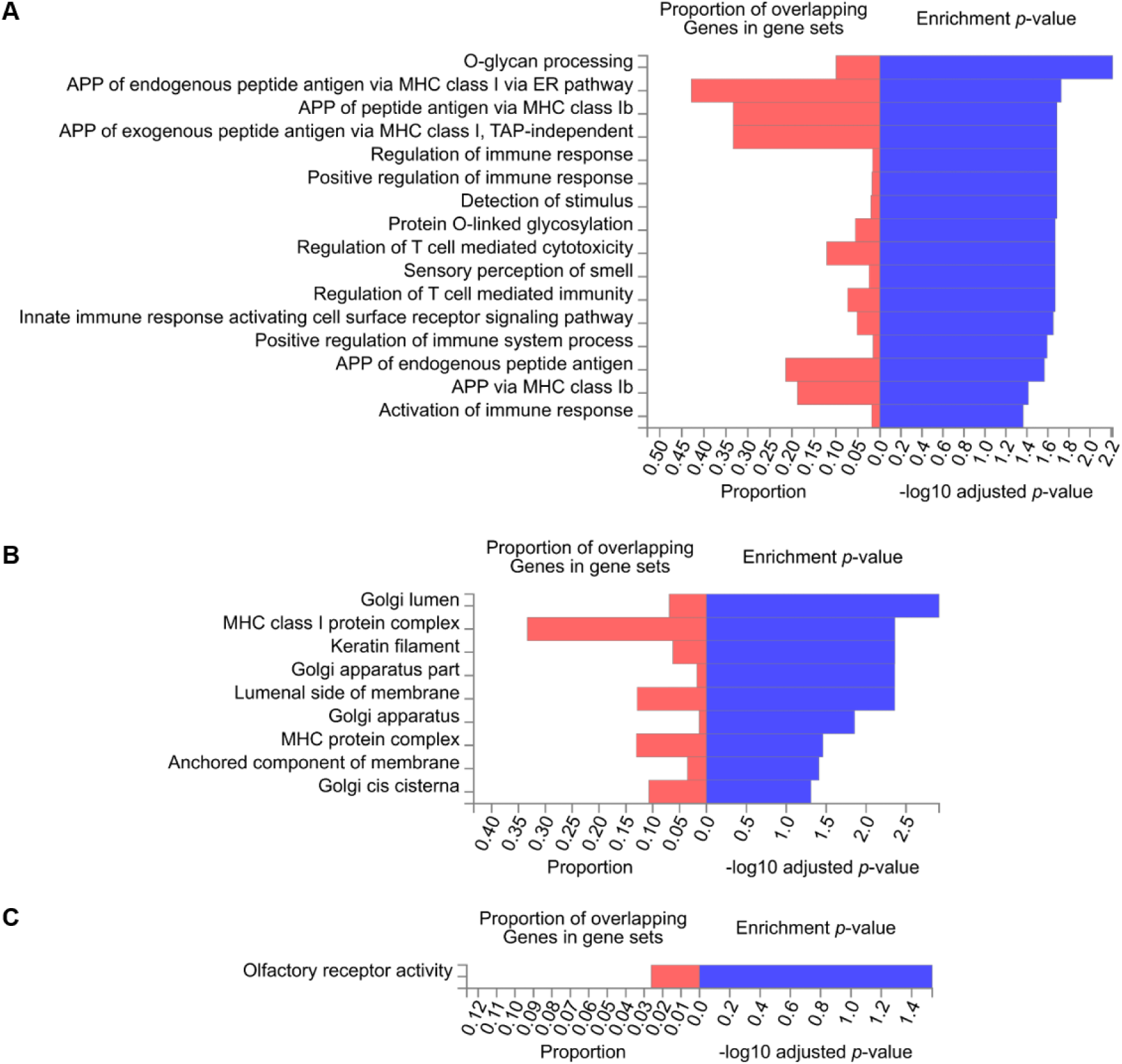
Overrepresentation analysis of genes containing variants with potential clinical relevance in the unaffected twin. Hypergeometric tests performed between the set of 227 genes that contains DVMTs of the unaffected twin, and the Gene Ontology (GO) gene sets for biological processes (BP), cellular components (CC) or molecular functions (MF) obtained from MsigDB database (https://www.gsea-msigdb.org/gsea/msigdb/), point to 16 overrepresented BPs **(A)**, 9 CCs **(B)**, and 1 overrepresented MF **(C)**. Red bars indicate the proportion of genes tested (elements of the set of 227 genes) for each GO predefined gene set. Benjamini-Hochberg (FDR) multiple test correction method for enrichment testing was used, and gene sets with adjusted *p*-value <0.05 were considered significative. The larger blue bars indicate that the test results are more likely to be non-random.

The affected twin showed 55 overrepresented BPs, 18 CCs, and 10 MFs **(Figures 4 and 5)**. The affected twin had the same overrepresented BPs, and CCs but with a higher proportion (∼25-55% and ∼45%, respectively) **(Figures 4 and 5A)**. “Serotonin-gated cation-selective channel activity” (GO:0022850) was the most overrepresented MF in the affected twin (∼60%) **(Figure 5B)**.

**Figure 4.**
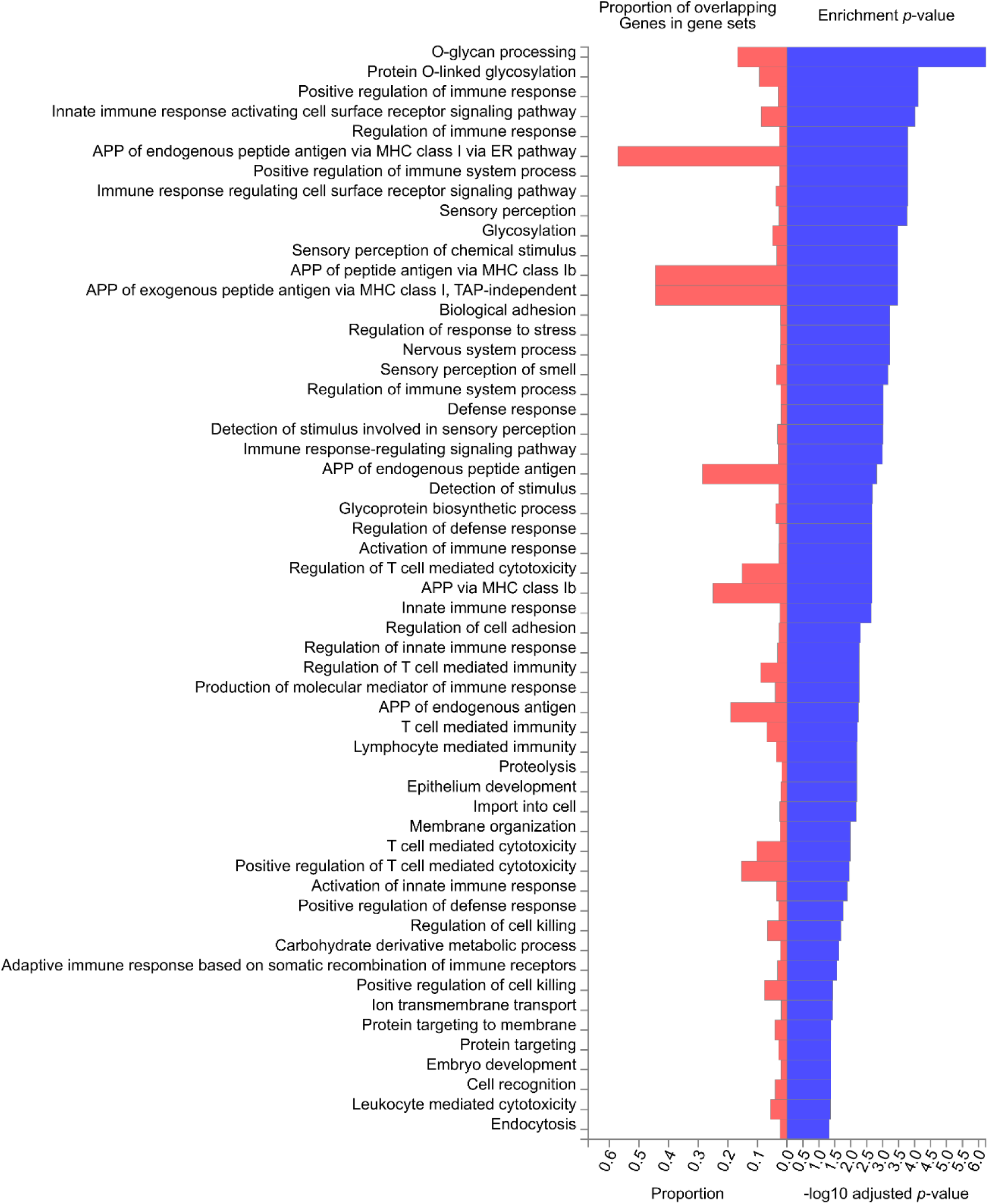
Overrepresentation analysis of genes containing variants with potential clinical relevance in the affected twin – biological processes. Hypergeometric tests performed between the set of 362 genes that contains DVMTs of the affected twin, and the Gene Ontology (GO) gene sets for biological processes (BP) obtained from MsigDB database (https://www.gsea-msigdb.org/gsea/msigdb/), point to 55 overrepresented BPs. Red bars indicate the proportion of genes tested (elements of the set of 362 genes) for each GO predefined gene set. Benjamini-Hochberg (FDR) multiple test correction method for enrichment testing was used, and gene sets with adjusted *p*-value <0.05 were considered significative. The larger blue bars indicate that the test results are more likely to be non-random.

**Figure 5.**
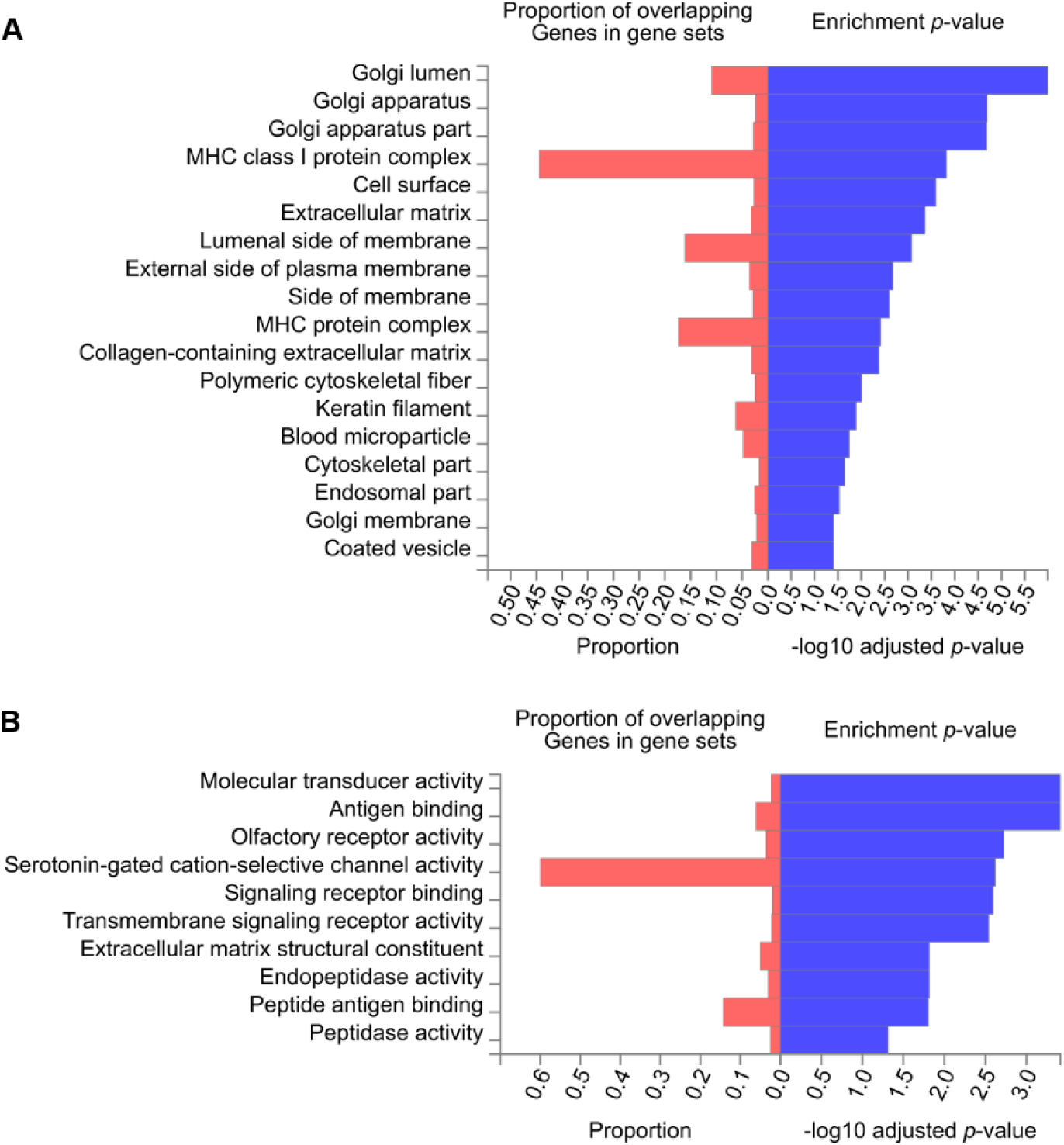
Overrepresentation analysis of genes containing variants with potential clinical relevance in the affected twin – cellular components or molecular functions. Hypergeometric tests performed between the set of 362 genes that contains DVMTs of the affected twin, and the Gene Ontology (GO) gene sets for cellular components (CC) or molecular functions (MF) obtained from MsigDB database (https://www.gsea-msigdb.org/gsea/msigdb/), point to 18 CCs and 10 overrepresented MFs. Red bars indicate the proportion of genes tested (elements of the set of 362 genes) for each GO predefined gene set. Benjamini-Hochberg (FDR) multiple test correction method for enrichment testing was used, and gene sets with adjusted *p*-value <0.05 were considered significative. The larger blue bars indicate that the test results are more likely to be non-random.

Regarding UPSIT, the narcolepsy affected twin had severe hyposmia (30/40) and the non-affected twin had mild hyposmia (10/40).

## Discussion

The whole-genome analysis of discordant twins for NT1 revealed that the twin pair here considered has identical HLA alleles distribution. However, several variants in genes outside the HLA, such as *MUC17, MUC12, GOLGA6L2*, and *FOXO3B* were observed (Supplementary Tables S3 and S4). Also, the unaffected twin has significantly more frameshift mutations as compared to the affected twin (108 versus 75) and mutations that affect stop codons (61 versus 5 in stop gain, 26 versus 2 in start lost) (Figure 2). The overrepresentation analysis of genes containing variants with potential clinical relevance in the unaffected twin reveals that most mutations are in genes related to immune regulation function, Golgi apparatus, MHC, and olfactory receptor. Because the HLA alleles associated with NT1 are present in both twins we hypothesize that the affected twin follows the expected phenotypic course while in the unaffected twin the mutations here identified might play a role in protecting the unaffected twin from NT1. The detailed analysis of each individual mutation, and eventually the experimental proof of cause-consequence is a future development of this work. Another perspective is the study of distribution and frequencies of the non-HLA mutations, here characterized, in broader cohorts.

## Data Availability

All sequence reads and BAM files here produced will be available upon request after this manuscript is accepted for publication.

